# AI-based analysis of the shunt treatment in pre- and post-surgery computed tomography brain scans of iNPH patients

**DOI:** 10.1101/2023.06.28.23292008

**Authors:** S. Shailja, Christopher Nguyen, Krithika Thanigaivelan, Vikram Bhagavatula, Jefferson W. Chen, B. S. Manjunath

## Abstract

**Background:** This study examines whether quantifiable changes can be detected in ventricular volume in Idiopathic Normal Pressure Hydrocephalus (iNPH) patients that undergo ventriculo-peritoneal shunt procedures. There is no known metric that characterizes the change in ventricular volume for iNPH patients after shunt placement.

**Methods:** Two de-identified and independent datasets are studied:

- 45 brain CT scans (24 diagnosed with iNPH and 21 normal elderly individuals) are used to evaluate the effectiveness of our proposed ventricular volume metric as a diagnostic tool for iNPH. The performance of our deep learning model-based metric is compared to the traditional Evan’s Index using ROC analysis.
- 16 subjects with a total of 50 longitudinal CT scans taken before and after shunt surgery across different imaging centers are studied to quantify the impact of shunt treatment. Clinical symptoms of gait, balance, cognition, and bladder continence are studied with respect to the proposed metric.

**Results:** Our proposed metric achieves high accuracy (0.95), precision (0.96), and recall (0.96) in distinguishing between normal and iNPH subjects, surpassing the performance of the Evan’s Index. This metric allows us to track changes in ventricular volume before and after shunt surgery for 16 subjects. Notably, the 15 subjects with iNPH demonstrate a decrease in ventricular volume post-surgery and a concurrent clinical improvement in their iNPH symptomatology.

**Conclusion:** Our novel metric accurately quantifies changes in ventricular volume before and after shunt surgery for iNPH patients, serving as an effective radiographic marker for a functioning shunt in a patient with iNPH.

- What is already known on this topic – The diagnosis of iNPH involves both clinical and radiographic stigmata. Radiologists rely largely on visual examination of CT scans and provide qualitative evaluations about ventricular volume.
- What this study adds – Our study provides quantitative information about the patency and function of the shunt.
- How this study might affect research, practice, or policy – The validated deep learning-based metric enhances iNPH diagnosis accuracy by tracking radiographic biomarkers. This facilitates decision-making regarding the efficacy of shunt surgery and the effect on brain compliance. We provide a web interface to apply the metric, its reliable performance across multiple institutional scanner types could be adapted to the real-time clinical evaluation of iNPH and improve treatment workflows.

## 1 INTRODUCTION

Idiopathic Normal Pressure Hydrocephalus (iNPH) is a neurodegenerative condition characterized by the accumulation of cerebrospinal fluid (CSF) in the brain, which results in ventriculomegaly. An estimate of more than 700,000 Americans have iNPH, but less than 20% receive an appropriate diagnosis (1). The typical symptoms of iNPH are gait disturbance, urinary incontinence, and progressive dementia (2).

Shunt surgery is an effective treatment for iNPH which diverts CSF from the ventricles to the peritoneal cavity (2). The surgically treatable nature of iNPH highlights the recent advancements in diagnostics and treatments that have improved patient outcomes (3). Follow-up studies of patients have shown positive results in gait followed by bladder and cognitive improvements (4). Symptoms are classified as improved if they result in an improvement in the patient’s day-to-day functioning, as assessed by the examiners and patients’ family members. However, this improvement is difficult to quantify and accurately track post-surgery because the brains are not compliant and the decrease in ventricular size is minimal. This highlights the need for a reliable method to assess the efficacy of shunt surgery for iNPH.

In recent years, radiological measures have been used to diagnose and quantify the impact of shunt surgery on patients with iNPH. The periventricular gray-white matter ratio on CT reveals high diagnostic accuracy in distinguishing iNPH cases from controls, particularly in the anterior periventricular area (5). The clinical improvement after shunt surgery in patients with iNPH is shown to be associated with a slight reduction of ventricular size (6). The authors found that the callosal angle and ventricular volume significantly increased and decreased, respectively, but the changes were too small to visually detect. A reduced callosal angle post-operatively might indicate under-drainage of CSF but no correlation between radiological measures and clinical improvement was found (7). Although the above-mentioned methods are manual or semi-automatic, they suggest that further research is required to improve the accuracy of quantifying CSF in ventricles as a radiographic marker. In this study, we propose a fully automated method with a user-friendly web interface for computing the ventricular volume metric, enabling the diagnosis and tracking of iNPH.

Volumetric analysis has emerged as a valuable tool for understanding various aspects of iNPH. The utilization of regional gray matter volume analysis and machine learning has proven effective in distinguishing responders from non-responders to temporary CSF drainage in iNPH, facilitating patient selection and outcome prediction (8). Furthermore, the evaluation of processes and outcomes in iNPH patients treated within the Adult Hydrocephalus Clinical Research Network revealed significant improvements following CSF drainage and shunt surgery, accompanied by a low rate of complications (9). The metric proposed in this paper holds promise for application in such investigations.

Artificial intelligence (AI) has been used for automatic ventricle segmentation for brain CT scans and MRIs. Prior research has developed AI-based tools (10, 11) to segment cerebral ventricles from T2-weighted MRIs and calculate ventricular volume in pediatric patients with obstructive hydrocephalus. In this paper,

- We propose a novel AI-based metric derived from ventricular volume in brain CT scans, offering a promising approach for diagnosing iNPH.
- We validate the efficacy of this metric by tracking ventricular volume changes after shunt placement using an independent dataset, providing compelling evidence of its applicability.
- Finally, we provide a user-friendly web interface that automates the computation of volumetric metrics for convenient analysis.

Our approach facilitates neurosurgical decision-making and provides valuable volumetric data on the ventricles for use in clinical settings. Figure 1 shows the computation of the proposed center ventricular volume metric and outlines the pipeline for utilizing the metric to differentiate iNPH patients from normal subjects and quantify ventricular volume changes post-shunt surgery.

**Figure 1:**
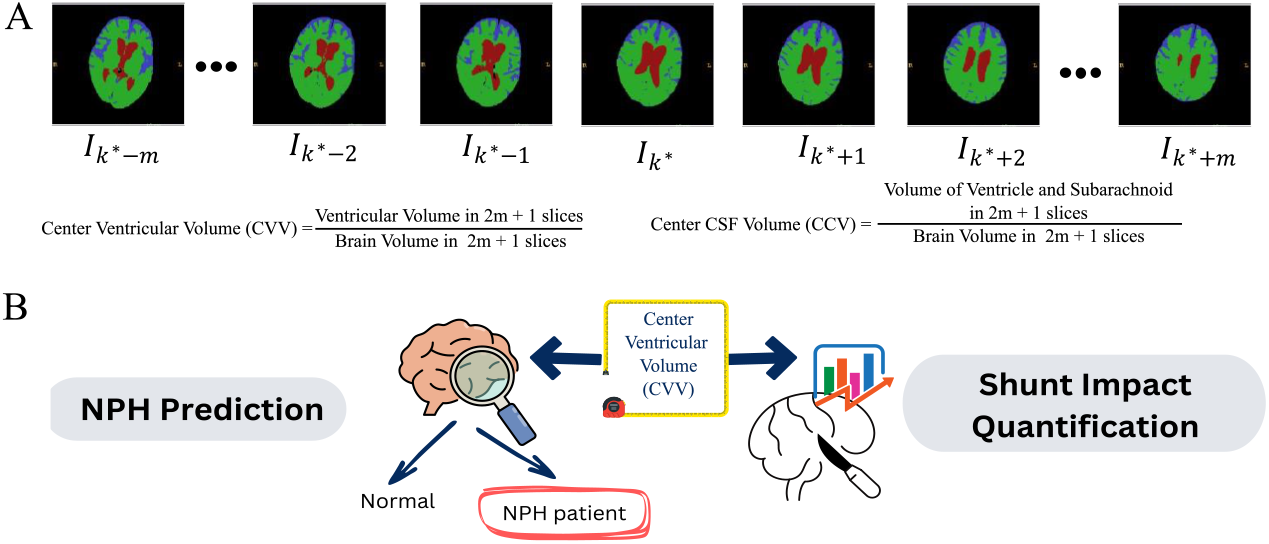
Computation of the center ventricular volume (CVV) and its usage. (A) CVV is computed as the ratio of ventricular volume in 2*m* + 1 slices to the brain volume in those 2*m* + 1 slices using the segmented scans. m slices are chosen before and after slice k* with the maximum ventricular volume. (B) CVV is used to distinguish between normal and iNPH subjects and to quantify the effect of shunt treatment by analyzing the segmented scans before and after surgery.

## 2 METHODS

### 2.1 Retrospective review of study populations

In this study, a total of 95 brain CT scans were acquired as part of the treatment process. Of these, 45 scans from 45 subjects (24 iNPH patients and 21 normal elderly control) are utilized to demonstrate the diagnostic value of the method in distinguishing iNPH from normal control subjects. The subjects were aged between 60s and 90s (mean=75; SD=7.7). The CT scans of adult patients who had a clinical and radiological diagnosis of iNPH from January 2015 to December 2022 are selected from the Neurosurgery NPH clinic run by author JWC at University of California Irvine (UCI) Medical Center. This retrospective study is conducted with all images de-identified by the UCI Center for Artificial Intelligence in Diagnostic Medicine (CAIDM) as specified by the IRB agreement between UCI Medical Center and the University of California, Santa Barbara (UCSB).

Additionally, 50 scans from 16 patients (8 males and 8 females) that had undergone a ventriculo-peritoneal (VP) shunt ranging in age from 60s to 90s (mean=79; SD=5.9) are utilized to compare the pre-operative and post-operative ventricular volumes. At least one pre-operative CT scan of the brain was taken. Multiple post-surgery scans were taken in addition to the immediate post-operative scan. The additional scans were taken as part of routine care usually after a change in the shunt programming. This is to ensure that there was no development of a subdural hematoma or subdural hygroma. Additional scans were also done if the patient had complaints such as headaches. Patients’ responses to shunting were obtained from retrospective chart reviews. Improvement in any of the clinical indices of iNPH (gait, cognition, urinary function) was given a score of +1. Worsening of symptoms was scored as -1. No change was scored as 0. If the patient had a normal gait, cognition, or urinary function before or after the shunt surgery, it was still scored 0. The distribution of effects on symptoms after surgery is shown in Table 1.

**Table 1:**
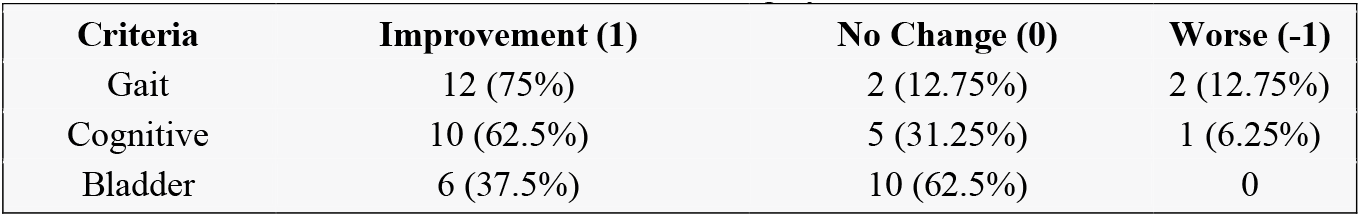
Distribution of subjects based on gait, cognitive, and bladder function improvements after shunt surgery.

Because of referral patterns and insurance restraints, CT scans were performed on different machines, which included scanners at UCI as well as scanners from other radiology centers. As CT scans are reported in different orientations (coronal, sagittal, axial), the images selected for our study are those in the axial-oblique orientation. These scans in DICOM format are converted to NIFTI for our analysis(12).

### 2.2 Ventriculoperitoneal shunt surgery

Ventriculoperitoneal shunts were placed via a burr hole in the right frontal region at Kocher’s point. The shunt tubing was tunneled posteriorly with a post-auricular intervening incision to travel across the clavicle and the abdomen where it was placed intraperitoneally using a general surgery laparoscopic technique. The Catheter was placed into the frontal horn using neuronavigation (Medtronics-Axiem EM system, Minneapolis, Minnesota). The shunt valve was placed in a subcutaneous pocket posterior to the ventricular entry site. The shunt used was the Codman-Certas programmable valve with Bactoseal proximal and distal catheters (Integra life sciences, Princeton, New Jersey). A post-operative CT scan of the head was done within 6 hours of the surgery to confirm the placement of the catheter and to ensure that there were no immediate intracranial complications. Patients were mobilized and the majority (90%) were discharged on the first post-operative day.

### 2.3 Review: Evan’s Index for NPH

Evan’s Index (EI) is a measure used in the diagnosis of iNPH (13). Evan’s Indices for this study were calculated using the Visage 7 (Visage Imaging, Inc., San Diego, CA, United States) system. Only images that did not have major artifacts such as from motion or excess noise were included in this study. EI is calculated by dividing the maximum width of the frontal horns of the lateral ventricles by the maximal parietal diameter of the brain at the same level on an axial CT scan. Radiologists often use EI as an indicator of enlarged ventricular volume, which can be a sign of NPH. The standard guidelines in the United States and internationally for iNPH indicate ventriculomegaly as an EI greater than 0.3 (14). However, there are several disadvantages to relying on EI for diagnosis:

- It is manually calculated, hence, prone to errors and variations in measurement depending on the skill of the person performing the calculation.
- The definitions of maximum width and parietal diameter are subjective, which can also affect the accuracy of the measurement.
- EI only reflects one slice of the 3D CT scan, so it may not provide a complete picture of the effect of iNPH on ventricular volume.

Furthermore, the reduction in ventricular volume due to shunt surgery can be difficult to observe on CT scans, and EI may not accurately capture this. To address these limitations, we develop an automated pipeline for diagnosing iNPH that is more accurate and objective in evaluating the impact of shunt placement post-surgery.

### 2.4 Automated pipeline for diagnosing NPH

#### 2.4.1 Segmentation of CT scan

A crucial step towards automated methods for the diagnosis of iNPH is brain image segmentation, which involves separating the brain into different regions of interest: ventricle, subarachnoid, and gray/white matter. In this study, we use a UNet-based network which is a peer reviewed method developed by Vision Research Lab at UCSB (11, 15) to segment brain CT scans.

#### 2.4.2 Center Ventricular Volume (CVV) metric

We propose a new metric, the Center Ventricular Volume to effectively quantify the ventricular volume, which is a key feature affected in NPH. CVV is based on the segmentation of brain scans that allows for the precise identification and quantification of regions of interest. We demonstrate the usage of the CVV in two ways:

- Distinguishing between normal and iNPH subjects -- The CVV can aid in an early identification of individuals at risk for the development of iNPH.
- Quantifying and tracking the effect of shunt treatment by analyzing the segmented scans before and after surgery.

By utilizing the CVV, we provide a more accurate and reliable method for assessing ventricular volume in iNPH patients. We provide a user-friendly pipeline that has the potential to improve the diagnosis and treatment of this condition.

To compute the CVV, consider the set of all 2D cross-sections (slices) of the brain in a scan, *I*, where each slice *k ∈ I* represents a specific cross-sectional view of the brain. Then, *k*^*∗*^ is the slice of the scan with the maximum ventricular volume that can be represented as

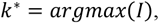

where *argmax(I)* is the argument that maximizes the function *I*, which in this case is the slice with the highest ventricular volume in the scan. The volume of any region (ventricle, CSF, brain) is calculated by multiplying the total number of voxels labeled as that region in the segmented scan by the voxel resolution. We have empirically chosen to analyze a constant brain thickness of *T*^*∗*^ = 35 *mm*. To determine the optimal slices for analysis, we first identify the slice *k*^*∗*^ with the maximum ventricular area. Then, we determine the number of slices required to achieve a thickness of *T*^*∗*^ = 35 *mm*, taking into account different thickness resolution of the scanned images. Denoting the number of slices above and below *k*^*∗*^ as *m*, we calculate *m* as follows:

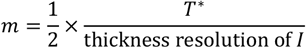

Hence, the selected slices for analysis are: *k, k* + 1, …, *k* + *m, k* − 1, *k* − 2, …, *k* − *m*. We then calculate the total ventricular volume (*V*_total_) and total brain volume (*B*_total_) in these slices. We define the CVV as:

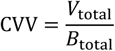

The CVV accounts for the variability in the image thickness resolution and provides a robust volumetric metric for assessing ventricular volume. *V*_total_ represents around 80% of the ventricular volume and 10% of the entire brain volume. Increase in CVV implies increase in ventricular volume. Table 2 and Table 3 present the computed metrics for each CT scan, demonstrating that CVV is a reliable representation of the brain ventricles allowing for differentiation between iNPH patients and normal individuals and quantification of improvement after shunt surgery. Similarly, we calculate the total CSF volume (*C*_total_) in these slices and the corresponding Center CSF Volume (CCV) metric as shown in Figure 1:

**Table 2:** Comparison of methods distinguishing iNPH subjects from Normal controls.

| Anonymized Sub id | Diagnosis | Age range | EI: Evan's Index | CVV: Center Ventricular Volume (Semi) | CCV: Center CSF Volume (Semi) | CVV: Center Ventricular Volume (UNet) | CCV: Center CSF Volume (UNet) |
| --- | --- | --- | --- | --- | --- | --- | --- |
| 1 | Norm | 80s | 0.28 | 0.06 | 0.16 | 0.08 | 0.16 |
| 2 | Norm | 90s | 0.33 | 0.08 | 0.24 | 0.08 | 0.21 |
| 3 | Norm | 60s | 0.22 | 0.03 | 0.08 | 0.03 | 0.05 |
| 4 | Norm | 70s | 0.31 | 0.08 | 0.20 | 0.08 | 0.16 |
| 5 | Norm | 70s | 0.32 | 0.04 | 0.13 | 0.05 | 0.14 |
| 6 | Norm | 60s | 0.31 | 0.09 | 0.25 | 0.12 | 0.21 |
| 7 | Norm | 60s | 0.26 | 0.04 | 0.16 | 0.04 | 0.11 |
| 8 | Norm | 60s | 0.26 | 0.04 | 0.15 | 0.04 | 0.11 |
| 9 | Norm | 60s | 0.26 | 0.05 | 0.15 | 0.06 | 0.12 |
| 10 | Norm | 70s | 0.27 | 0.06 | 0.16 | 0.06 | 0.15 |
| 11 | Norm | 70s | 0.27 | 0.09 | 0.26 | 0.08 | 0.19 |
| 12 | Norm | 70s | 0.29 | 0.08 | 0.30 | 0.07 | 0.20 |
| 13 | Norm | 70s | 0.32 | 0.10 | 0.20 | 0.09 | 0.17 |
| 14 | Norm | 70s | 0.29 | 0.05 | 0.17 | 0.05 | 0.10 |
| 15 | Norm | 60s | 0.26 | 0.04 | 0.18 | 0.05 | 0.17 |
| 16 | Norm | 70s | 0.28 | 0.06 | 0.25 | 0.07 | 0.18 |
| 17 | Norm | 70s | 0.27 | 0.06 | 0.14 | 0.06 | 0.12 |
| 18 | Norm | 70s | 0.28 | 0.10 | 0.17 | 0.10 | 0.16 |
| 19 | Norm | 80s | 0.31 | 0.12 | 0.23 | 0.10 | 0.16 |
| 20 | Norm | 60s | 0.30 | 0.05 | 0.15 | 0.05 | 0.13 |
| 21 | Norm | 60s | 0.29 | 0.04 | 0.11 | 0.04 | 0.10 |
| 22 | NPH | 70s | 0.39 | 0.23 | 0.28 | 0.23 | 0.26 |
| 23 | NPH | 70s | 0.39 | 0.22 | 0.29 | 0.22 | 0.29 |
| 24 | NPH | 80s | 0.33 | 0.12 | 0.29 | 0.13 | 0.28 |
| 25 | NPH | 80s | 0.32 | 0.11 | 0.26 | 0.12 | 0.25 |
| 26 | NPH | 80s | 0.31 | 0.10 | 0.27 | 0.10 | 0.26 |
| 27 | NPH | 80s | 0.33 | 0.11 | 0.30 | 0.11 | 0.27 |
| 28 | NPH | 80s | 0.38 | 0.19 | 0.31 | 0.19 | 0.31 |
| 29 | NPH | 70s | 0.37 | 0.14 | 0.32 | 0.14 | 0.29 |
| 30 | NPH | 70s | 0.37 | 0.14 | 0.31 | 0.14 | 0.28 |
| 31 | NPH | 80s | 0.36 | 0.10 | 0.28 | 0.11 | 0.27 |
| 32 | NPH | 80s | 0.33 | 0.08 | 0.26 | 0.09 | 0.27 |
| 33 | NPH | 80s | 0.35 | 0.14 | 0.27 | 0.14 | 0.24 |
| 34 | NPH | 70s | 0.36 | 0.19 | 0.27 | 0.20 | 0.26 |
| 35 | NPH | 80s | 0.37 | 0.15 | 0.25 | 0.14 | 0.20 |
| 36 | NPH | 80s | 0.39 | 0.14 | 0.23 | 0.15 | 0.22 |
| 37 | NPH | 80s | 0.39 | 0.16 | 0.26 | 0.16 | 0.23 |
| 38 | NPH | 70s | 0.37 | 0.17 | 0.27 | 0.17 | 0.27 |
| 39 | NPH | 70s | 0.36 | 0.17 | 0.26 | 0.17 | 0.24 |
| 40 | NPH | 70s | 0.43 | 0.19 | 0.28 | 0.19 | 0.25 |
| 41 | NPH | 70s | 0.38 | 0.16 | 0.29 | 0.16 | 0.27 |
| 42 | NPH | 70s | 0.36 | 0.16 | 0.27 | 0.18 | 0.29 |
| 43 | NPH | 70s | 0.38 | 0.20 | 0.31 | 0.19 | 0.29 |
| 44 | NPH | 70s | 0.38 | 0.20 | 0.30 | 0.22 | 0.30 |
| 45 | NPH | 80s | 0.40 | 0.19 | 0.31 | 0.19 | 0.31 |

**Table 3:** Computation of Evan’s Index, CVV metric, and difference in ventricular volume in cc for subjects with pre- and post-surgery CT scans.

| Anonymized Sub_id | Diagnosis | Shunt surgery | EI: Evan's Index | Center Ventricular Volume (CVV) | Change in Center Ventricle Volume (cc) (Pre – post) |
| --- | --- | --- | --- | --- | --- |
| 1 | worsening of cog fxn (could be attributed to covid or fall) | Pre | 0.352 | 0.21 |  |
|  | improved gait function, improved cognition has reverted to pre-op levels | Post | 0.349 | 0.2 | 3 |
| 2 | Freezing of gait at start, urinary urgency, mild cog impairment | Pre | 0.35 | 0.22 |  |
|  | continued gait problems + bradykinesia | Post | 0.314 | 0.18 | 23 |
| 3 | gait issues progressed to ataxic, slow, unbalanced; some memory/cog issues | Pre | 0.345 | 0.22 |  |
|  | gait has much improved | Post | 0.32 | 0.18 | 18 |
| 4 | progressively worsening gait | Pre | 0.421 | 0.26 |  |
|  | memory+thinking improved; gait also improved | Post | 0.398 | 0.24 | 9 |
| 5 | slow gait, sway side to side; some memory deficit | Pre | 0.384 | 0.18 |  |
|  | feeling of gait improvement, more fluent speech | Post | 0.376 | 0.17 | 10 |
| 6 | wheelchair bound at this time, incontinent | Pre | 0.43 | 0.17 |  |
|  | some/slow improvement | Post | 0.428 | 0.16 | 2 |
| 7 | <restricted access to medical record> | Pre | 0.319 | 0.12 |  |
|  | Improved bladder, overall improvement | Post | 0.298 | 0.09 | 15 |
| 8 | fall | Pre | 0.294 | 0.17 |  |
|  | another fall, slightly ataxic but fluid | Post | 0.292 | 0.18 | 4 |
| 9 | worsening gait; urinary urgency/incontinence | Pre | 0.334 | 0.22 |  |
|  | no more incontinence | Post | 0.31 | 0.17 | 29 |
| 10 | magnetic gait | Pre | 0.338 | 0.21 |  |
|  | able to walk independently, still magnetic/ataxic gait; improved gait fxn | Post | 0.326 | 0.16 | 30 |
| 11 | wide-based, slow, ataxic gait | Pre | 0.33 | 0.22 |  |
|  | improved gait, cognitive and urinary fxn are the same | Post | 0.32 | 0.18 | 21 |
| 12 | cognitive/memory impairment; urinary incontinence | Pre | 0.33 | 0.2 |  |
|  | made gain with gait and continence; cognitive has not increased or decreased | Post | 0.307 | 0.16 | 27 |
| 13 | progressive gait disturbance since 2015, severe balance problems; increased urinary frequency; memory is being impacted | Pre | 0.33 | 0.196 |  |
|  | still ataxic and wide-based, but can walk without walker; slight improvement with continence | Post | 0.316 | 0.199 | -2 |
|  | still ataxic and wide-based, but is fluid; shunt adjusted from 4 to 3 | Post (after shunt adjustment) | 0.308 | 0.18 | 10 |
| 14 | some memory impairment; some urinary incontinence | Pre | 0.306 | 0.14 |  |
|  | fall; still has incontinence; shunt level 4 | Post | 0.298 | 0.16 | -9 |
|  | slow, but fluid gait, not magnetic, another fall; worsening memory; shunt adjusted from 4.5 to 5 | Post (after shunt adjustment) | 0.3 | 0.11 | 29 |
| 15 | gait difficulty; urinary incontinence; mild memory loss | Pre | 0.306 | 0.17 |  |
|  | no improvement in incontinence; can stand for 2 mins but not walk yet; codman certa shunt level 5 to 4 | Post | 0.303 | 0.2 | -20 |
|  | can take small steps but gait is still ataxic and slow; memory is stable; uses adult briefs; current shunt setting is two | Post (after shunt adjustment) | 0.3 | 0.15 | 33 |
| 16 | slow, wide-based gait + ataxic; incontinence, | Pre | 0.43 | 0.24 |  |
|  | some improvement in gait, continence greatly improved | Post | 0.42 | 0.22 | 6 |

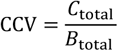

CCV can also aid in distinguishing between iNPH and normal control as shown in Table 2, but it is less sensitive in tracking the progression of iNPH or quantifying the effectiveness of shunt surgery (see Supplementary Table 1).

## 3 RESULTS

We compute the CVV metric for the first dataset that consists of 45 CT scans. The metric is calculated for both semi-automated segmented scans (automatically segmented using UNet and corrected by experts) and fully automated UNet trained on corrected CT scans. Additionally, EI is manually computed for all the scans. To classify the scans into iNPH and Normal, we compute the threshold of each metric and method using a Receiver Operating Characteristic (ROC) curve (16). The ROC curve allows us to evaluate the performance of each metric and method by plotting the true positive rate against the false positive rate at various threshold values. By examining the curve, we determine the optimal threshold that balances sensitivity and specificity for each metric and method. The threshold for EI is 0.3 which is also standard threshold to determine the iNPH condition (17). Table 4 displays the computed threshold and provides accuracy, precision, and recall metrics for each method. Our results indicate that the CVV and CCV metrics perform best for the classification task as shown in Figure 2. Table 2 contains the CVV and CCV metric computation for each subject using all the methods.

**Table 4:** Comparison of methods to distinguish between Normal vs NPH.

|  | Methods | Threshold | Accuracy | Precision | Recall |
| --- | --- | --- | --- | --- | --- |
| Manual | Evan's Index | 0.3 | 0.86 | 0.79 | 1 |
| Semi-automated (Semi) | Center Ventricular Volume Metric | 0.1 | 0.95 | 0.96 | 0.96 |
|  | Center CSF Volume Metric | 0.3 | 0.93 | 1 | 0.87 |
| Fully automated (UNet) | Center Ventricular Volume Metric | 0.1 | 0.95 | 0.96 | 0.96 |
|  | Center CSF Volume Metric | 0.2 | 0.98 | 1 | 0.96 |

**Figure 2:**
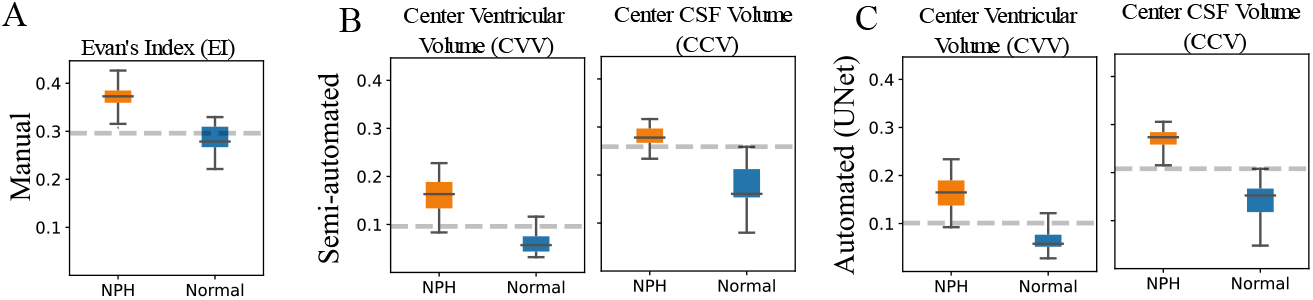
Comparison of metrics computed from different methods. (A) Evan’s Index, traditionally used to distinguish between iNPH and Normal, is manual, subjective, and based on a 2D slice. (B) CVV and CCV are computed on 3D volume of the UNet method’s automated initial segmentation, which is then manually corrected by radiologists. (C) CVV and CCV are computed on 3D volume of the fully automated UNet segmented brain CT scans.

The second dataset consists of a total of 50 CT scans from pre- and post-surgery scans of 16 patients. We compute the CVV for all the scans and compute the ventricular volume (*V*_total_). A clear trend towards a decrease in ventricular volume and CVV in post-surgery scans is observed that significantly correlates with the qualitative improvement in the patient’s iNPH condition as shown in Table 3. All subjects with iNPH showed a positive response to shunt surgery.

Figure 3A shows the subjects with scans collected within 3 months of surgery. For the patient with sub_id = 8 in Table 3, CVV did not decrease. This patient did not have iNPH but had post-traumatic hydrocephalus by history when he was referred for evaluation of ventriculomegaly 12 years after the initial severe traumatic brain injury which explains why CVV was not sensitive to the shunt. In Figure 3B, we show the subjects with scans collected after 3 months with time differences between scans ranging up to 40 months. This shows the longer-term impact of shunt treatment and the quantitative validation of this improvement by the CVV metric. It is known that the response to shunt surgery for iNPH is commonly perceived to be of limited duration, but we show here that even after 3 months, the ventricular volume is lower than pre-surgery scans. Figure 3C consists of subjects (sub_id = 13, 14, 15 in Table 3) who do not positively respond to the first shunt surgery. After additional time and shunt adjustments to augment CSF flow, subsequent CT scans show a decrease in the ventricular volume. This highlights the importance of the continued clinical evaluation and shunt adjustments to optimize the efficacy of the shunt. The ventricular volume computed after the first surgery shows no response or deterioration after shunt surgery. This is accurately captured and quantified with our method in the CVV computation. So, the metric effectively helps neurosurgeons in key decision-making regarding shunt valve adjustment. This enables more objective comparison of scans from a radiological point of view. The decrease in ventricular volume after shunt surgery also correlates with gait, cognition, and bladder improvement in the subjects as shown in Figure 3D. Also, we observe that the iNPH condition deteriorates before surgery resulting in increase in CVV metric with time until surgery.

**Figure 3:**
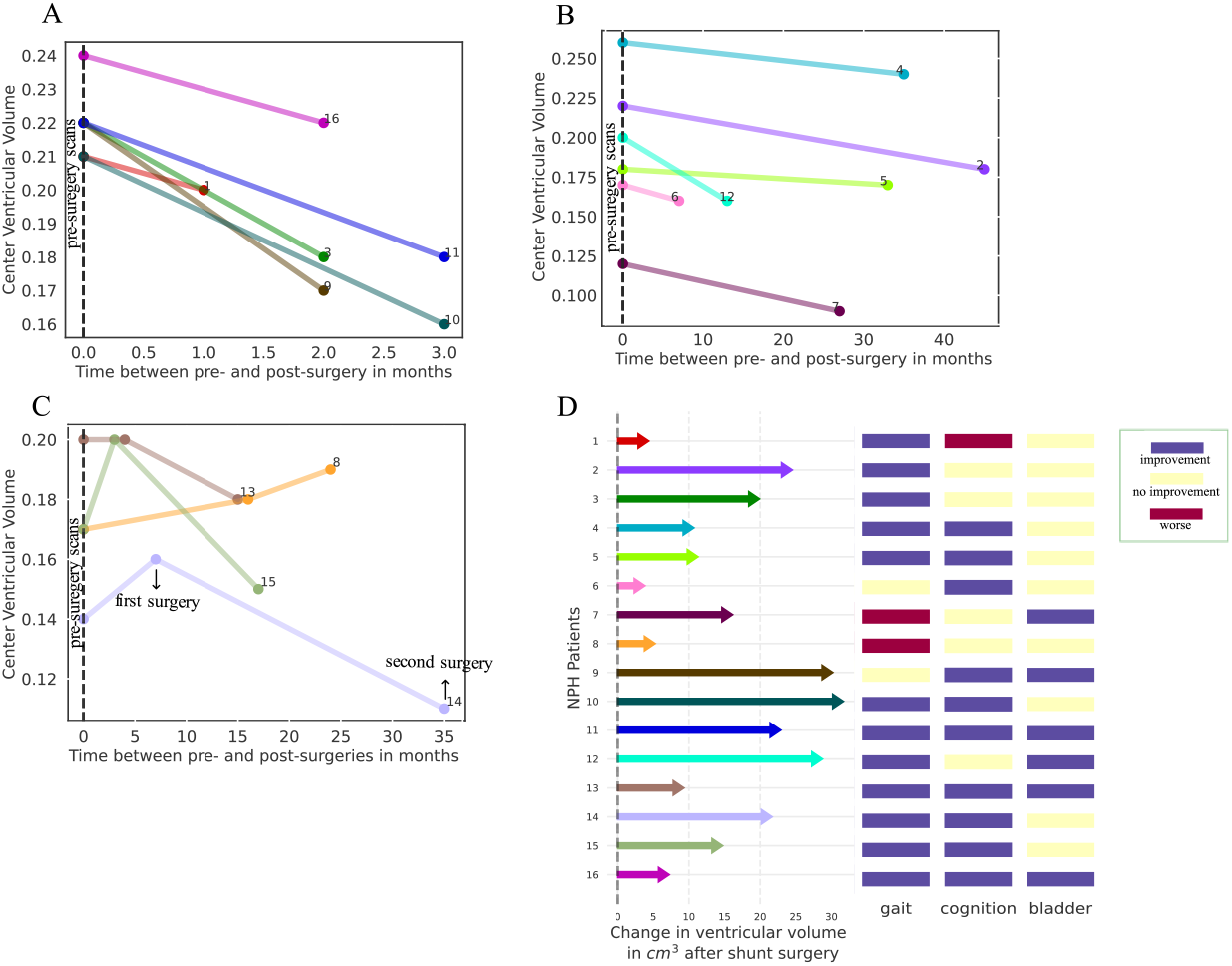
Decrease in ventricular volume after shunt surgery. (A) Pre- and post-surgery ventricular volume within three months of the shunt surgery. (B) Pre- and post-surgery ventricular volume after three months of the shunt surgery for distinct set of subjects. (C) Ventricular volume reduction in iNPH patients not responding to the first shunt surgery, including one patient (in yellow) who did not respond to the shunt placement for post-traumatic hydrocephalus. (D) Absolute change in CSF for ventricular volume in cc after shunt surgery for all the subjects, showing the correlation between improvement in patients and changes in ventricular volume.

## 4 DISCUSSION

### 4.1 Demonstration of decreased ventricular volume as a reliable indicator of shunt function

Our proposed method has the potential to serve as a tool to access shunt function (Figure 4). The advantages over other techniques include its non-invasive nature and the ease of obtaining a CT scan of the head. Periodic CT scans are done as part of routine care to ensure that patients do not develop subdural hematomas or subdural hygromas. Periodic quantitative evaluation of the ventricular volume can provide assurance of continued shunt functioning. We have not established the threshold on decrease in ventricular volume from pre-op status in cc that translates to improve or adequate drainage of CSF via the VP shunt. This is an area of future research.

**Figure 4:**
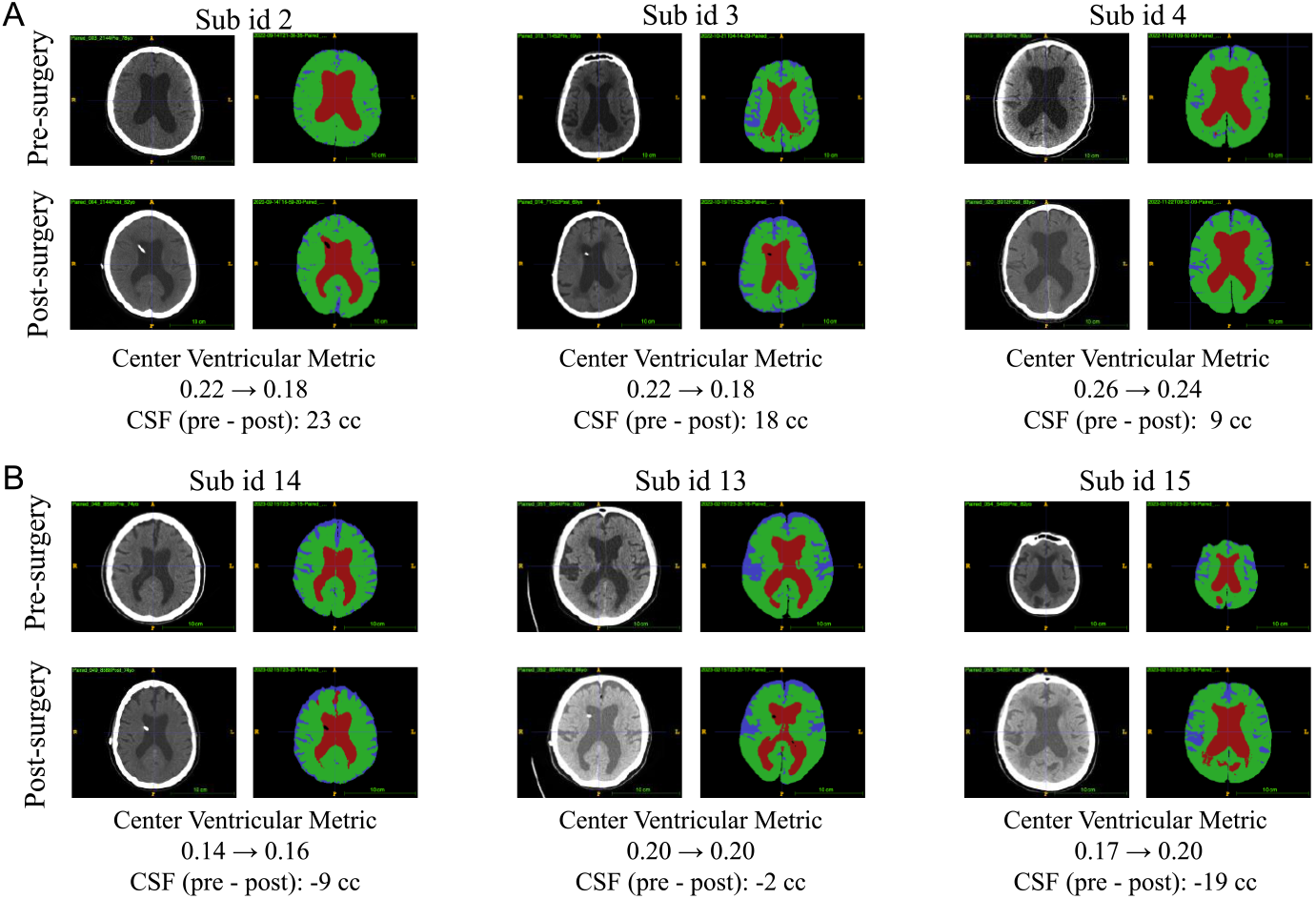
Ventricular volume analysis for iNPH patients. (A) Decrease in CVV and ventricular volume shows positive response to shunt surgery. (B) Increase in CVV and ventricular volume indicating worsening iNPH conditions. The iNPH shunt valve was adjusted for these patients. There is decrease in the CVV and absolute ventricular volume after second shunt surgery in non-responders to the first surgery. This demonstrates the potential utility of the ventricular volume metric in determining the need for shunt adjustment. It can be challenging to accurately evaluate the condition based solely on CT scan visualization.

### 4.2 Evan’s Index fails to reliably track the response to shunt surgery

In our analysis, we utilize Evan’s Index (EI) to monitor changes before and after shunt surgery in all subjects from the second dataset. Although EI demonstrates values greater than 0.3 for all iNPH patients, it fails to accurately correlate with the response to treatment, particularly in slow responders where shunt effectiveness was observed to be limited based on clinical diagnosis and subsequent adjustments to shunt diameter. Three patients (sub_id = 13, 14, 15 in Table 3) were slow responders to shunt treatment and required additional shunt programming adjustments. After the initial surgery, we observe that ventricular volume increased by 2, 9, and 20 cc respectively. EI falsely indicates a decrease in index values from 0.33 to 0.316, 0.306 to 0.298, and 0.306 to 0.303, respectively. This misleading trend could suggest that the shunt was effective without requiring further investigation. In contrast, our proposed metric demonstrates a consistent increase: 0.196 to 0.199, 0.14 to 0.16, and 0.17 to 0.2. These findings align with the clinical diagnosis, which indicated deteriorating gait, balance, cognition, and bladder continence. This provides strong evidence of the value of our metric in accurately tracking the impact of shunt surgery and optimizing CSF egress via the shunt. Following the shunt re-programming, we observe a decrease in both CVV and absolute volume, as expected, further validating the effectiveness of our metric in monitoring changes post-shunt surgery.

### 4.3 Significance of CSF ventricular quantification

CT scans of the head are routinely performed on iNPH patients with VP shunts because they are quickly done and provide anatomical information that neurosurgeons may use in their treatment paradigms. One of the known complications of over-shunting in iNPH patients is the development of subdural hematoma or subdural hygroma, especially after a fall. The advantage of a CT scan over an MRI is its rapid acquisition and the lack of shunt magnetic artifact that occurs with the MRI. The ability to demonstrate a decrease in ventricular volume is significant for demonstrating that there is patency and functioning of the VP shunt. The clinical significance is that with NPH, the lack of improvement or the deterioration in symptoms may not necessarily be from shunt dysfunction. Demonstration that the decrease in ventricular volume is maintained directs the clinician to look for alternative explanations.

### 4.4 Implications for brain compliance

The lack of visible decrease in the ventricular size after VP shunting has traditionally been attributed to a decreased brain compliance in iNPH (18). There is a myriad of age-related factors that affect brain compliance including arterial and venous calcifications and aqueductal narrowing or stenosis. This is one of the first demonstrations that the ventricular volume and size may change with shunting and correlate with improvement. This suggests that the CSF egress via the shunt may affect the overall brain compliance.

### 4.5 Universal nature of CT ventricular quantification

In this study, the CT scans that were used were derived from different CT scanners at different institutions. This is the nature of the care of these patients. Often patients have the CT scans done at their home institute because of proximity or insurance restraints. We were able to convert these DICOM images and obtain meaningful quantitative evaluations. Additionally, it should be noted that the axial-oblique orientation was selected for the analysis to provide some consistency of measurement. The total volume achieved in the center slices used in this study was chosen to encompass the areas that were most representative (80%) of the ventricular volume of the brain. However, it clearly is not the entire brain volume (10%). Specifically, the posterior fossa and 4^th^ ventricle are not included. This was intentional, as these areas are hard to capture consistently on the axial CT scans and there are some variations.

### 4.6 Limitations

The study may be limited by the absence of patients with fixed valves who required shunt removal due to serious over drainage symptoms. Our patient selection was biased as we selectively included patients who showed improvement after shunt surgery. Additionally, as iNPH typically affects elderly patients with co-morbidities such as Parkinson’s or Alzheimer’s disease, the findings may not be generalizable to those populations. One post-operative scan was selected for shunt impact comparison, while more CT scans over time may provide additional information about the progression of the changes in the ventricular volume. This is the focus of a future study.

## 5 CONCLUSIONS

The timely detection and screening of patients with iNPH is crucial as surgical interventions can effectively alleviate or reverse the symptoms. Our study utilized an AI tool to provide consistent and objective measurement of cerebral ventricular volume over time for iNPH patients. The proposed ventricular metric shows reliability across various institutional scanners and variations in slice thickness. It is computationally fast (1-2 minutes for brain segmentation and less than 10 seconds for volumetric computation on average) and has the potential to serve as a valuable clinical adjunct in standardizing and enhancing patient care. All data relevant to the study, including the brain CT scans used in this study will be publicly available through Bisque (19). The methods used in the study are available online through the UCSB Bisque web-based portal (20) and the code is available on GitHub (21, 22).

## Data Availability

All data produced in the present study will be made available after peer-review.

## 5 ACKNOWLEDGEMENTS

This research was supported by NSF award: SSI # 1664172. We acknowledge the support of the Bisque team at UCSB and the Center for Artificial Intelligence in Diagnostic Medicine (CAIDM) team at UCI for their assistance with data management.

---

**Supplementary Table 1:**
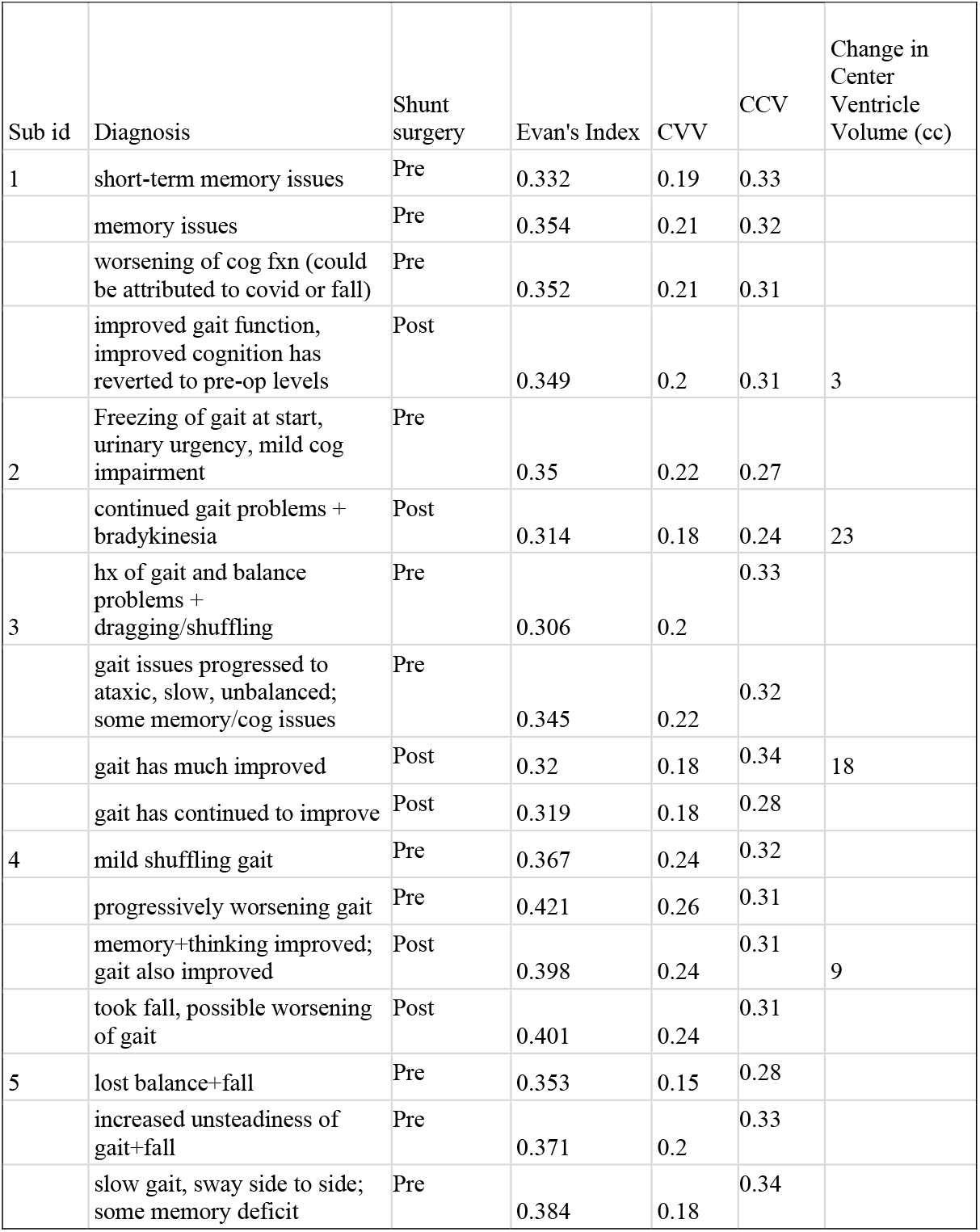
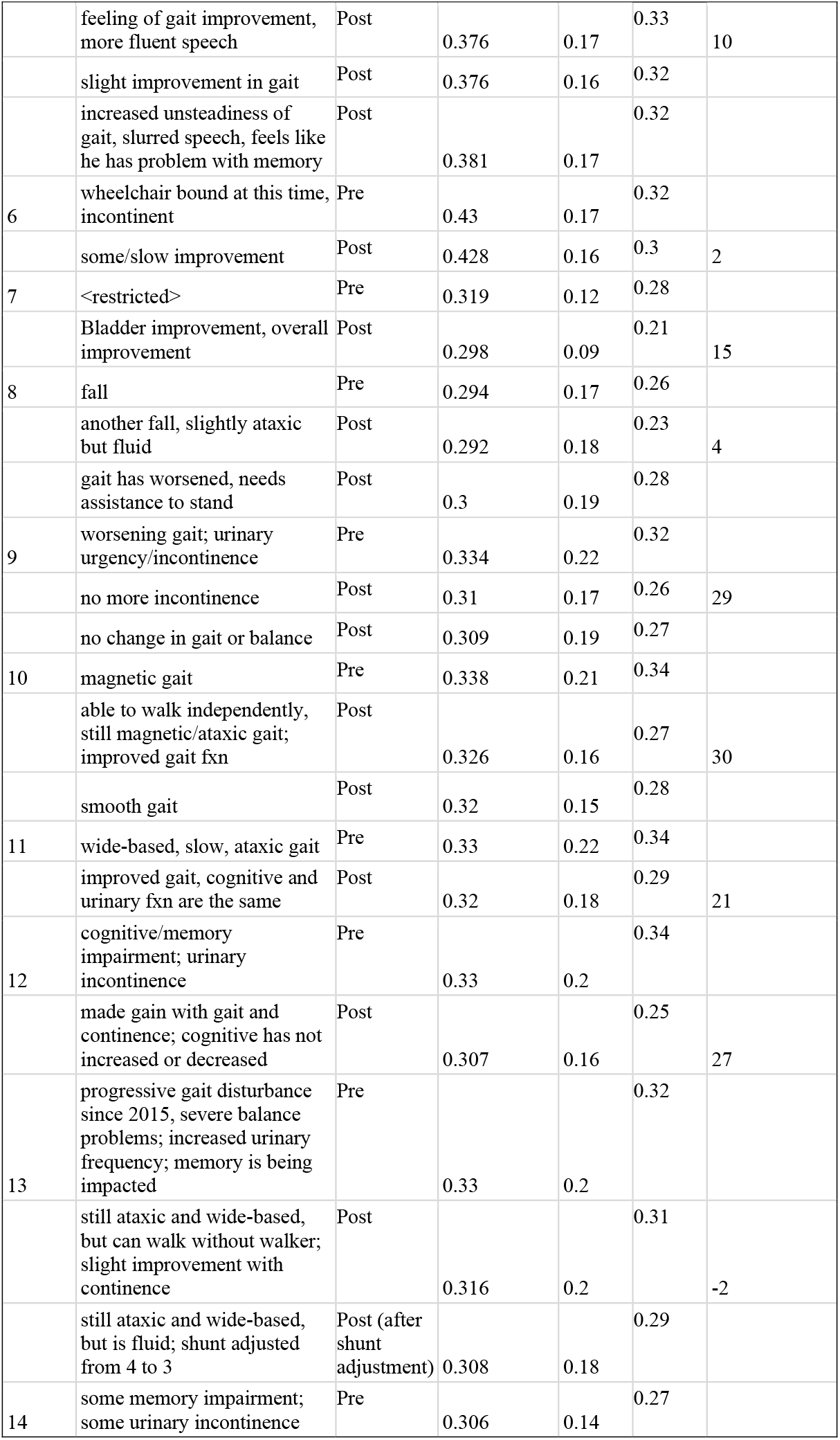
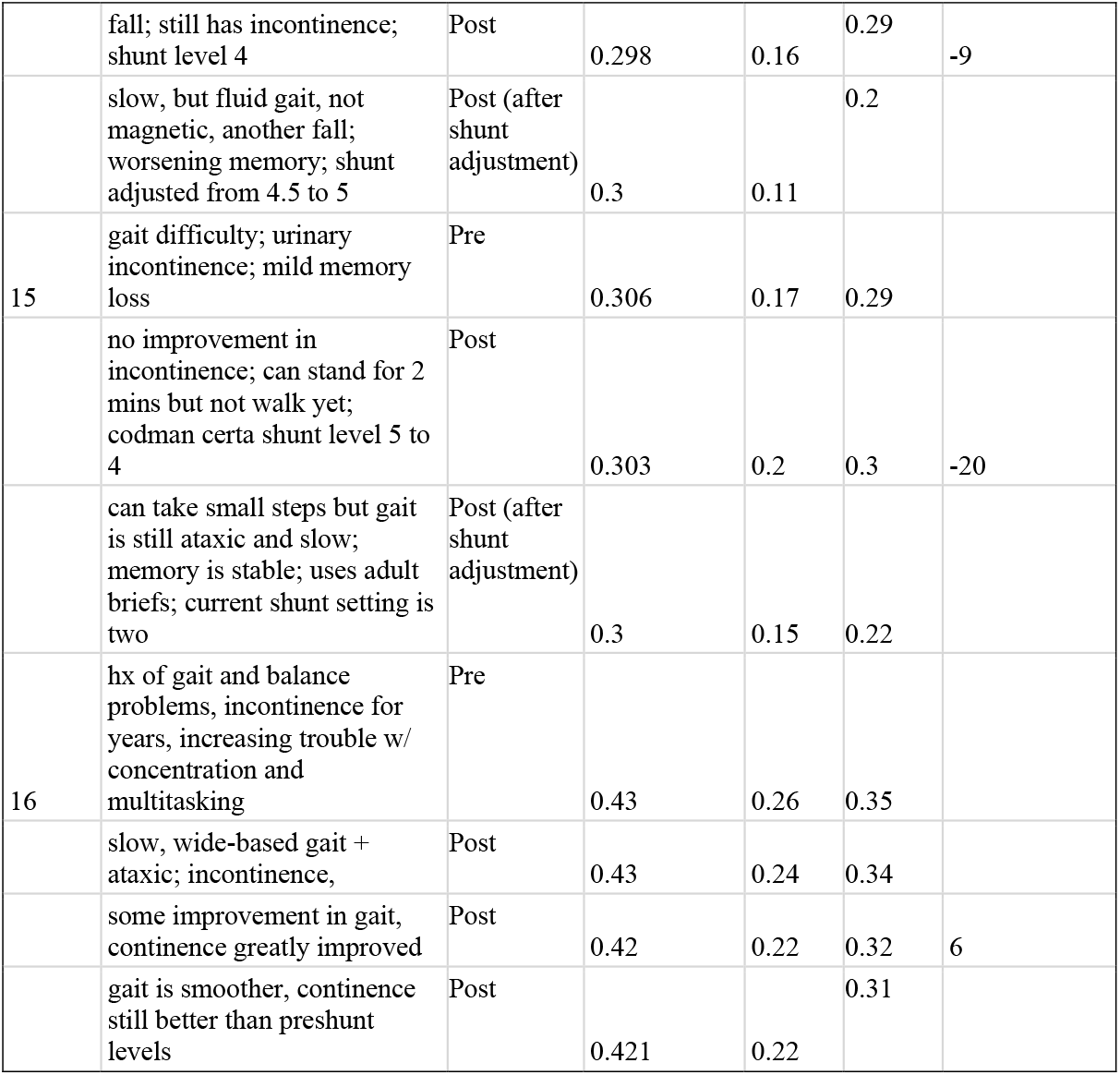
Computation of Evan’s Index, CVV metric, and ventricular volume in cc for subjects with pre- and post-surgery CT scans.

